# Probability of elimination for COVID-19 in Aotearoa New Zealand

**DOI:** 10.1101/2020.08.10.20172361

**Authors:** Rachelle N. Binny, Shaun C. Hendy, Alex James, Audrey Lustig, Michael J. Plank, Nicholas Steyn

## Abstract

On 25^th^ March 2020, New Zealand implemented stringent lockdown measures (Alert Level 4, in a four-level alert system) with the goal of eliminating community transmission of COVID-19. Once new cases are no longer detected over consecutive days, the probability of elimination is an important measure for informing decisions on when certain COVID-19 restrictions should be relaxed. Our model of COVID-19 spread in New Zealand estimates that after 2-3 weeks of no new reported cases, there is a 95% probability that COVID-19 has been eliminated. We assessed the sensitivity of this estimate to varying model parameters, in particular to different likelihoods of detection of clinical cases and different levels of control effectiveness. Under an optimistic scenario with high detection of clinical cases, a 95% probability of elimination is achieved after 10 consecutive days with no new reported cases, while under a more pessimistic scenario with low case detection it is achieved after 22 days.

## Introduction

On 28th February 2020, the first case of COVID-19 arrived in New Zealand and by 25th March the outbreak had reached 205 (confirmed and probable) cases (Ministry of Health, 2020). At this time, stringent lockdown restrictions (Alert Level 4, in a four-level alert system) were implemented with the goal of eliminating community transmission (Baker et al, 2020). Here, we define elimination as there being no active cases that could contribute to future community transmission. This definition excludes cases that are no longer infectious or that are in government-managed isolation and quarantine facilities.

Four weeks of Alert Level 4 restrictions were effective at reducing New Zealand’s effective reproduction number to 0.35 (Binny et al, 2020) and numbers of reported cases declined to less than 10 new cases per day. From 27th April (11.59pm), there was a slight easing of certain lockdown restrictions (Alert Level 3) (for example, schools – years 1 to 10 – and Early Childhood Education centres were permitted to open with limited capacity) which remained in place for a further three weeks. Following these 7 weeks of stringent restrictions, daily numbers of new cases had dropped to between zero and one.

In order to declare elimination of COVID-19, it is useful to know how likely it is that the virus has actually been eliminated after consecutive days of zero new reported cases, or conversely, the likelihood of there being undetected community transmission. The stochastic model of Plank et al (2020) can be simulated to estimate the probability, *P(elim)*, of having eliminated COVID-19 in NZ after a given number of consecutive days with no new reported cases. Such estimates are important to inform decisions on timings for the easing of certain COVID-19 restrictions. A high probability of elimination, for example *P(elim)* greater than 0.95, means there is only a small risk of cases that remain undetected in NZ; however, so long as the probability of elimination is less than one, there is still a chance of undetected cases sparking a new outbreak. Here, we report the probability of elimination under different scenarios of case detection and control effectiveness in NZ.

## Results

Under an optimistic scenario with high detection and reporting of clinical cases and moderate effectiveness of Alert Levels 2-3, our model estimates *P(elim)=0.95* after 10 consecutive days with no reported cases (Fig. 1). For a pessimistic scenario with low detection of clinical cases and low effectiveness of Alert Levels 2 and 3, *P(elim)=0.95* is achieved after 22 days with no reported cases (Fig. 1).

**Figure 1:**
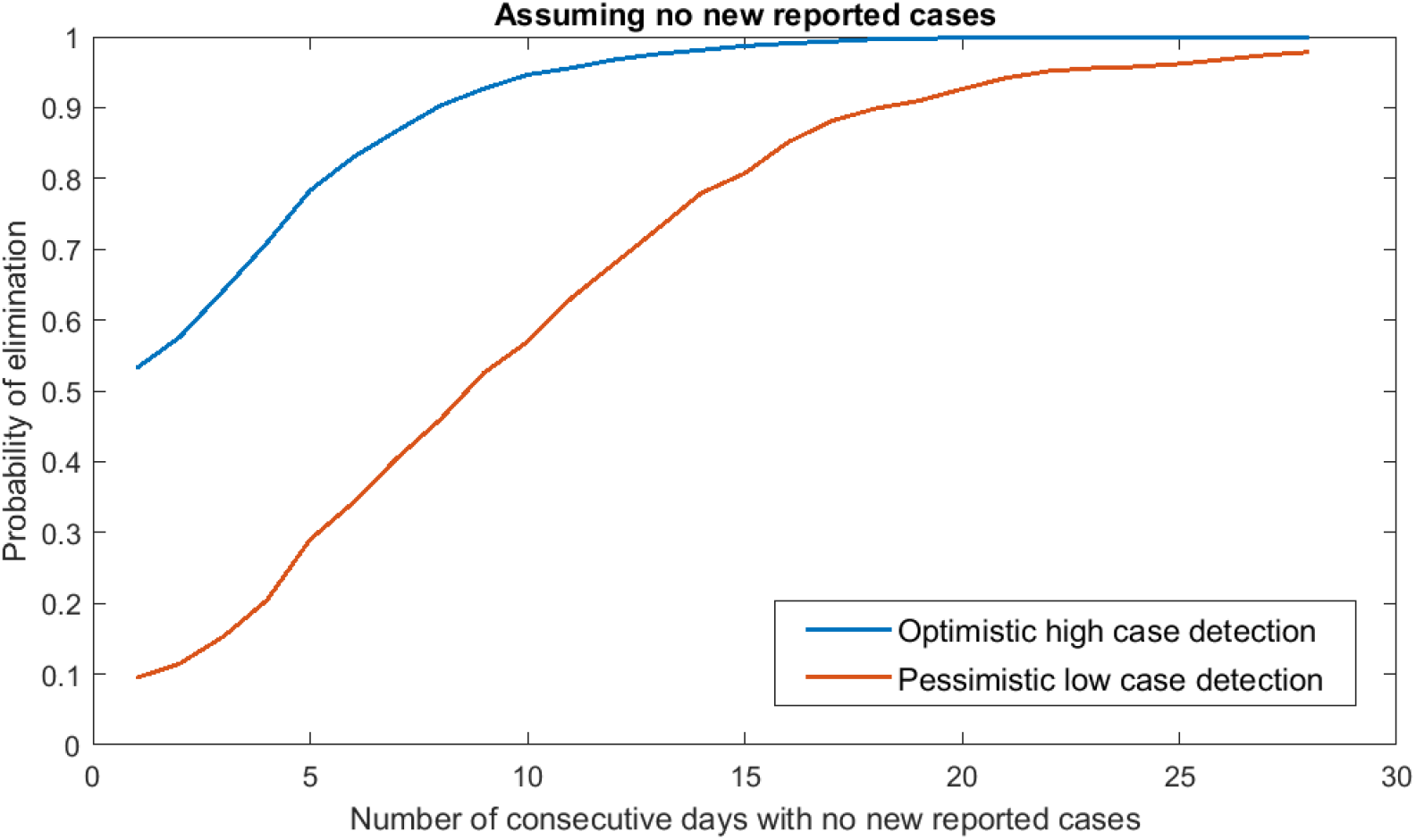
Probability of elimination, P(elim), after different numbers of consecutive days with no new reported cases. Optimistic scenario with high detection and reporting of clinical cases (p_R_=75%) and moderate effectiveness of Alert Levels 2 (C(t)=0.75, Reff=1.8) and 3 (C(t)=0.4, Reff=0.95). Pessimistic scenario with low detection and reporting of clinical cases (pR=20%) and low effectiveness of Alert Levels 2 (C(t)=0.95, Reff=2.3) and 3 (C(t)=0.9, Reff=2.2).

### Sensitivity analyses

We assessed the extent to which probability of elimination varied under different proportions of clinical cases that are detected and reported, *p_R_*, different distributions of generation times, and for different relative transmission rates, *C(t)*, for Alert Levels 2 and 3. For sensitivity analyses we used the shorter reporting delay employed in Plank et al (2020) (distribution from isolation to reporting, r (shape = 1, scale = 3.48); following Price et al (2020) and estimation using limited NZ data). For the scenarios above, using this shorter reporting delay gave similar results to those in Fig. 1, with *P(elim)=*0.95 achieved after 12 consecutive days with no reported cases in the optimistic scenario (Table 1) and after 22 days in the pessimistic scenario (results not shown).

**Table 1:** Sensitivity of P(elim) to varying proportion of clinical cases detected and reported, Pr. Relative transmission rate C(t)=1, 0.75, 0.4 and 0.15 for Alert Levels 1-4, respectively.

| Proportion of clinical cases detected and reported | $P_R=20\%$ (low detection) | $P_R=50\%$ | $P_R=75\%$ | $P_R=90\%$ (high detection) |
| --- | --- | --- | --- | --- |
| $R_{\text{eff}}$ when $C(t)=1$ | 2.47 | 2.42 | 2.37 | 2.35 |
| Days until $P(\text{elim})=0.95$ | 22 | 14 | 12 | 10 |

Results were moderately sensitive to changing the proportion of clinical cases that are detected and reported, *p_R_* (Table 1) and to the mean of the Weibull-distributed generation time (Table 2). For instance, with low detection and reporting of clinical cases (*p_R_*=20%), 22 consecutive days with no new reported cases are required to achieve *P(elim)*=0.95. When detection is very high (*p_R_*=90%), *P(elim)*=0.95 can be achieved after 10 days. Increasing the mean generation time from 5 days *(Wei(*scale = 5.67, shape = 2.83)) to 8 days (*Wei*(scale = 9.0, shape = 2.83)) increased the consecutive number of days required to achieve *P(elim)*=0.95 to 22 days. Results were relatively insensitive to different choices of relative transmission rate *C(t)* for Alert Levels 2 (Table 3) and 3 (Table 4).

**Table 2:** Sensitivity of P(elim) to varying the mean and variance of the Weibull-distributed generation time. Relative transmission rate C(t)=1, 0.75, 0.4 and 0.15 for Alert Levels 1-4, respectively. Proportion of clinical cases detected and reported, p_R_=75%.

| Generation time distribution | $\text{Wei}(\text{scale} = 5.67, \text{shape} = 2.83)$ | $\text{Wei}(\text{scale} = 9.0, \text{shape} = 2.83)$ | $\text{Wei}(\text{scale} = 5.67, \text{shape} = 1.5)$ |
| --- | --- | --- | --- |
| $R_{\text{eff}}$ when $C(t)=1$ | 2.37 | 2.21 | 2.36 |
| Days until $P(\text{elim})=0.95$ | 12 | 22 | 13 |

**Table 3:** Sensitivity of P(elim) to varying the relative transmission rate, C(t), of Alert level 3. Relative transmission rate C(t)=1, 0.75 and 0.15 (corresponding to Ref = 2.37, 1.78 and 0.35) for Alert Levels 1, 2 and 4, respectively. Proportion of clinical cases detected and reported, p_R_=75%.

| Alert Level 3<br>transmission | $C(t)=0.2,$<br>$R_{\text{eff}}=0.47$<br>(high<br>effectiveness) | $C(t)=0.3,$<br>$R_{\text{eff}}=0.72$ | $C(t)=0.4$<br>$R_{\text{eff}}=0.95$ | $C(t)=0.5,$<br>$R_{\text{eff}}=1.19$ | $C(t)=0.6,$<br>$R_{\text{eff}}=1.42$ | $C(t)=0.7,$<br>$R_{\text{eff}}=1.66$<br>(low<br>effectiveness) |
| --- | --- | --- | --- | --- | --- | --- |
| Days until<br>$P(\text{elim})=0.95$ | 9 | 12 | 12 | 12 | 12 | 12 |

**Table 4:** Sensitivity of P(elim) to varying the relative transmission rate, C(t), of Alert level 2. Relative transmission rate C(t)=1, 0.4 and 0.15 (corresponding to Ref = 2.37, 0.95 and 0.35) for Alert Levels 1, 3 and 4, respectively. Proportion of clinical cases detected and reported, p_R_=75%.

| Alert Level 2<br>transmission | $C(t)=0.6,$<br>$R_{\text{eff}}=1.42$ (high<br>effectiveness) | $C(t)=0.75$<br>$R_{\text{eff}}=1.78$ | $C(t)=0.9$<br>$R_{\text{eff}}=2.14$ | $C(t)=1,$<br>$R_{\text{eff}}=2.37$ (low<br>effectiveness) |
| --- | --- | --- | --- | --- |
| Days until $P(\text{elim})=0.95$ | 10 | 12 | 12 | 12 |

## Methods

A full description of the model is given in Plank et al (2020). Estimates were obtained by simulating 1000 realisations of the model and calculating probability of elimation, *P(elim)*, as the proportion of model realisations that achieve elimination (under our definition above) within a given timeframe. We used the parameter values given in Plank et al (2020) and James et al (2020), with a longer reporting delay of 6 days (distribution from isolation to reporting, Γ(shape = 1, scale = 6)) and the best-fit estimates for reproduction number *R_eff_* prior to and during Alert Level 4 reported in Binny et al (2020). For the optimistic scenario, we assumed 75% of clinical cases are detected and reported (*p_R_*=75%) and the transmission rate relative to no population-wide control, *C(t)*, was set to *C(t) = 1*, 0.75, 0.4 and 0.15 (corresponding to *R_eff_* = 2.37, 1.78, 0.95 and 0.35) for Alert Levels 1-4, respectively. The pessimistic scenario used a lower pr=20% and *C(t) =* 1, 0.95, 0.9 and 0.15 (corresponding to *R_eff_* = 2.47, 2.34, 2.22 and 0.36). The model was simulated up to 31 July 2020, using case data (sourced from Ministry of Health) up to 28th May 2020 and assuming that Alert Level 2 restrictions remain in place after that date.

## Discussion

Our model estimates that after 2 to 3 weeks of no new reported cases, there is a 95% probability that COVID-19 has been eliminated in NZ. Our findings were relatively robust to changes in model parameters; the number of consecutive days with no new reported cases required to achieve *P(elim*)=0.95 ranged from 9-22 days depending on assumptions about detection rate, control effectiveness and generation time. These estimates are slightly lower than the 27-33 days reported in a similar modelling study by Wilson et al (2020), obtained using a stochastic version of CovidSIM (http://covidsim.eu/).

Some important factors need to be considered when interpreting our results to inform decision-making. First, these estimates of probability of elimination only apply so long as there continues to be consecutive days with no reported cases, i.e. a new reported case resets the probability of elimination back to zero. Second, while the probability of elimination remains below one there is still a chance of undetected cases that could give rise to new outbreaks. The risk of an outbreak occurring undetected is likely to be higher for communities who experience inequitable access to primary healthcare and testing, including Māori and Pacific peoples (McLeod et al, 2020; James et al, 15 May 2020). Furthermore, so long as there remains a chance of undetected cases, the easing of restrictions (for instance removing limits on the size of gatherings) increases the risk of super-spreading events arising. Arrival of new COVID-19 cases at the border also presents an ongoing risk of future community transmission. These risks can be partly mitigated by continuing to implement testing and surveillance systems that are sufficient to detect community transmission, and by ensuring equitable access to healthcare and testing; by fast and effective contact tracing and case isolation (James et al, 12 June 2020); and by effective testing and quarantine for all new arrivals at the border.

## Data Availability

This article does not present new data.

